# Neurocognitive Functioning is Impaired in Perinatally HIV-Infected Youth

**DOI:** 10.1101/2024.08.28.24312647

**Authors:** Tamara Welikson, Manoj K. Sarma, Margaret Keller, James Sayre, Irwin Walot, David E. Michalik, Judy Hayes, Karin Nielsen-Saines, Jaime Deville, Andrea Kovacs, Eva Operskalski, Joseph A. Church, M. Albert Thomas, Joseph Ventura

## Abstract

**Background:** The present study examined neurocognitive differences between Perinatally HIV (PHIV)-infected-youth and age and gender matched healthy controls. Despite early, long-term anti-viral treatment (ART), significant neurocognitive deficiencies remain for PHIV-infected-youth reaching adulthood compared to controls.

**Methods:** Participants were assessed with a comprehensive neuropsychological battery. An Overall Neurocognitive Composite Score and a Global Deficit Score (GDS) were created. Sleep, depression, and developmental level of intellectual functioning were also examined.

**Results:** PHIV-youth performed more poorly than controls in all neurocognitive domains. Very large effect sizes were observed for the Overall Neurocognitive Composite Score and GDS. PHIV-infected-youth appear to be significantly more depressed compared to controls, but there were no differences in amount or type of sleep observed.

**Conclusion:** Despite early, long-term anti-viral treatment (ART), neurocognitive deficiencies remain for PHIV-infected-young-adults. The verbal learning domain was significantly impaired with implications for functioning. The PHIV-infected-youth were also depressed and not receiving treatment for depression.

## 1. Introduction

Perinatal Human Immunodeficiency Virus (PHIV)-infected youth appear associated with impairments in neurocognitive (Cohen et al., 2014; Nagarajan et al., 2012; Yadav et al., 2017), neurodevelopmental (Yadav et al., 2017), and psychiatric assessment performance (Chernoff et al., 2018; Willen et al., 2017). Global cognitive measures suggest that individuals diagnosed with PHIV perform below average in most cognitive domains including executive functioning, working memory, cognitive flexibility, motor performance, language, and speeded information processing (Cohen et al., 2014; Hermetet-Lindsay et al., 2017; Malee, Smith, & Mellins, 2016; Nagarajan et al., 2012; Robbins et al., 2019; Sirois et al., 2016; Willen et al., 2017). Compared to healthy controls, research suggests PHIV-infected-youth do not perform as well on measures of general cognition or visuospatial functioning (Laughton et al., 2013).

Imaging studies have identified neurological deficits and brain atrophy before the emergence of adolescence, when cognitive functioning, including memory, language, reading skills, processing speed, attention, concentration, executive control, planning, cognitive flexibility, problem-solving, and behavior rapidly develops (Laughton et al., 2013). Chronic exposure to a neurotropic infection during CNS development is a risk for PHIV (Smith & Wilkins, 2015). The more damage and dysregulation that occurs in the brain during illness onset, the more likely an individual’s cognitive reserve may be impacted (Basso & Bornstein, 2000). Although these young people continue to develop cognitively in the presence of illness (Ashby et al., 2015), they are subject to increased risk for developmental language impairment throughout adulthood (Laughton et al., 2013; Malee et al., 2016).

Research suggests PHIV-infected youth are at higher risk for psychiatric illness due to genomic, biological, environmental, psychosocial (Abrams et al., 2018; Laughton et al., 2013; Mellins & Malee, 2013; Smith & Wilkins, 2015), and cognitive issues (Hermetet-Lindsay et al., 2017; Laughton et al., 2013; Mellins & Malee, 2013) compared to undiagnosed individuals. Individuals with PHIV-infection often reported symptoms of depression, anxiety, behavioral difficulties, suicidal ideation/attempt, hyperactivity, attention difficulties, and sleep disturbance (Hermetet-Lindsay et al., 2017; Le Prevost et al., 2018; Malee et al., 2011; Mellins & Malee, 2013; Smith et al., 2019; Vreeman et al., 2015), which also result in high hospitalization rates for comorbid psychiatric disorders (Vreeman et al., 2015). Thus, depressive symptoms might impact developmental levels of functioning and neurocognitive development which might in turn impact functional outcomes into adulthood.

Evidence of neurodevelopmental, neurocognitive, and comorbid mental illness have been found in PHIV-infected youth. However, only a limited number of studies have used comprehensive assessment batteries that included measures of depression, sleep, neurocognitive deficits, and developmental level of intellectual functioning for PHIV-infected-youth with extensive treatment histories. Little is known about the long-term neurological effects of ART on brain development and functioning in PHIV-infected young adults (Laughton et al., 2013). Therefore, our study examined cognitive functioning and clinical features of viral pathogenesis and long-term early intervention ART treatment on PHIV-infected-youth.

A comprehensive neurocognitive battery was administered and included assessments of depression and sleep to assess the effects of these parameters on neurocognitive functioning. We studied young adults receiving long-term cART compared to age and gender matched healthy controls in an effort to better understand the long-term potential deficits of youth diagnosed with PHIV and to minimize potential confounding variables between groups. We hypothesized that PHIV-infected young adults would perform more poorly compared to healthy controls on most neurocognitive assessments and on assessments of mood and sleep. Also, we hypothesized that psychosocial impairments after a life-time of HIV infection would be substantial with many areas impacted by HIV and its treatment.

## 2. Methods

We report the findings from a study for which the primary aim was brain imaging in PHIV-infected-youth. As part of that study, a neurocognitive battery was administered using a cross-sectional, paired subject design.

### 2.1. Subject population and recruitment

A total of 22 participants were recruited and enrolled, including 11 PHIV-infected-youth who received early and prolonged antiretroviral treatment and 11 healthy control subjects. These PHIV-infected-youth who were receiving combination antiretroviral treatment (ages 18-30) were recruited from HIV clinics in the greater Los Angeles area: Los Angeles County Harbor-University of California Los Angeles (UCLA) Medical Center (Departments of Pediatrics and Medicine) in Torrance, Miller Children’s Hospital, Long Beach, David Geffen School of Medicine at UCLA, Department of Pediatrics, Los Angeles, and Los Angeles County/University of Southern California Medical Center, Department of Pediatrics, Los Angeles. Gender and age matched control subjects were individuals who responded to flyers on college campuses. Of the 11 healthy controls, seven were in college (6) or junior college (1) and 4 had completed a degree (AA, Bachelor’s or Master’s). Only 3 of the PHIV-infected-youth were in college or junior college and none had completed college.

There were 3 male and 8 female subjects in both the PHIV-infected-youth (patient) and matched paired control groups. The mean age of the patients was 22.5 years (+/- 2.9 years, range 19.6 years to 29.1 years) while the control subjects were 22.4 years (+/- 2.4 years, range 20.1 years to 28.4 years). The mean age difference between patient and matched control subjects was 0.9 years (+/- 0.5 years, range 0.2 years to 1.9 years).

#### 2.1.1 Inclusion criteria

Ages 18-30 years, perinatal HIV transmission if HIV-infected, currently receiving combination antiretroviral therapy if HIV-infected in follicular phase of menstrual cycle (to avoid hormonal influence on cerebral metabolites in an associated concurrent magnetic resonance spectroscopy study), negative urine pregnancy test, and HIV–1/2 OraQuick Advance Rapid Antibody Test negative for controls (testing was performed after consent obtained).

#### 2.1.2 Exclusion criteria

History of central nervous system disease, seizure disorder (controls only), severe metabolic disturbance (i.e. renal or hepatic failure), metal implants in the body or brain, claustrophobia, Attention Deficit Hyperactivity Disorder (ADHD), Substance Use Disorders, pregnancy, medications other than asthma medications (controls only), females in luteal phase of menstrual cycle, current clinical psychiatric care for psychiatric illness diagnoses, severe school difficulties (controls only), and Hepatitis C infection. Subjects were excluded if they met criteria for Unspecified Alcohol-Related Disorders (7 or more alcoholic drinks per week) and Cannabis-Related Disorders (any cannabis use within the past 6 months). All participants were non-smokers, except for one patient. One patient was found on subsequent chart review to have a significant problem with depression without history of psychiatric care.

Informed, signed consent was obtained from each subject using an Institutional Review Board (IRB) consent form approved by the Los Angeles Biomedical Research Institute at Los Angeles County Harbor-UCLA Medical Center (now Lundquist Institute). Both the Los Angeles Biomedical Research Institute and UCLA gave IRB approval which also included MRI/MRS imaging, neurocognitive assessment, and a detailed review of the patient subjects’ medical records (Sarma et al., 2021).

#### 2.1.3 Subject characteristics

Characteristics of the 11 PHIV-infected-youth and the age and gender paired controls are summarized in Table 1 including CD4 counts and viral load data close to the time of testing, and age of treatment for HIV. Testing for CD4 and viral load was within 72 days of the neuropsychological assessment, except for one patient for whom the elapsed time was three months. Although viral load was undetectable for 6 patients, the values (copies/mL) for the remaining subjects were 22,800, 1002, 884, 119,536, and 31. The patient with the highest viral load later advised her physician she had treatment adherence problems. We were unable to assess lifetime treatment adherence. Patients received HIV treatment at a young age (Table 1). Of the PHIV-infected-youth, 3 were African Americans and 8 were Caucasian (8 of Hispanic ethnicity). Of these 11 PHIV-infected-youth, 4 patients had HIV encephalopathy and one patient was thought to have a probable diagnosis of HIV encephalopathy. For control subjects, 5 identified as Caucasian (3 identified of Hispanic ethnicity), 5 identified as Asian and 1 identified as African American/Caucasian. All participants were right-handed with the exception of 2 patients who were primarily left-handed. Parental education (in years) data was available for 9 patients and 11 controls (Table 1). Of the 11 PHIV patients, 8 had a history of school difficulties, potentially demonstrating effects of HIV on the brain even in the absence of an HIV encephalopathy diagnosis. Of these 11 patients, 5 had a definite clinical CDC classification of C and one had a possible classification of C. One patient had been treated with Adderall for ADHD previously when in school. Medical records were reviewed to determine if there was evidence of substance abuse for the mother. Of the 11 mothers, there was evidence of substance abuse for 2 mothers and for one mother information was not available. For 8 mothers, there was no evidence of substance abuse. Information regarding maternal substance abuse was not available for paired control subjects.

**Table 1.** Evaluation of PHIV+ Patients (n=11) and Paired Healthy Controls (n=11) on Demographic and Viral Load Characteristics.

|  |  | PHIV+ Patients | Paired Healthy Controls | p-Value |
| --- | --- | --- | --- | --- |
| Age | Mean Age in Years (SD) | 22.5 (2.9) | 22.4 (2.4) | 0.93 |
|  | Mean Age Treatment was Initiated Months (SD) | 21.6 (37) | N/A |  |
|  | Age Range in Months Treatment was Initiated | 3-129 | N/A |  |
|  | % Patients Treated at Less Than One Year of Age | 64% | N/A |  |
| Sex | Female<br>Male | 73%<br>27% | 73%<br>27% | 1.00 |
| Education | Mean in Years (SD) | 12.09 (1.04) | 15.00 (1.41) | <0.001 |
| Parent Education* | Mean in Years (SD) | 11.50 (2.45) | 14.36 (2.54) | 0.07 |
| Race | Caucasian | 8 | 5 | 0.04 |
|  | African American | 3 | 1 | 0.32 |
|  | Asian | 0 | 5 | 0.25 |
| Ethnicity | Hispanic | 8 | 3 | 0.13 |
|  | Non-Hispanic | 3 | 8 | 0.13 |
| Viral Load | % viral load < 20 copies /mL | 55% | N/A | - |
|  | % viral load < 1500 copies/mL | 82% | N/A | - |
|  | Mean log<sub>10</sub> viral load (SD) | 2.2 (1.4) |  | - |
| CD4 Count | Mean (SD) Range | 507 (301) 55-963 | N/A<br>N/A | - |
|  | % CD4 count < 200 | 18% | N/A | - |

\* Not all participants provided parental education

### 2.2. Comprehensive Neuropsychological Assessment Battery

All subjects were administered a two-hour standardized, comprehensive neuropsychological assessment battery by a trained member of the project in accordance with assessment instructions in English. Neurocognitive Measures were grouped into 11 cognitive domains and 2 composite scores for analysis. These measures are summarized in Table 2. Raw data and Z-scores were transformed into t-scores by utilizing established normative data (Beck, Steer, & Garbin, 1988; Buysse et al., 1989; Carey et al., 2004; Nuechterlein et al., 2008; Rey & Osterrieth, 1993; Tombaugh, 2014; Wechsler, 2001). Executive Functioning, Psychomotor Functioning, and Abstract Thinking raw scores were calculated into t-scores based on the performance of controls (*N* = 11). Higher t-scores signified better performance across all measures.

**Table 2.** Comprehensive Neurocognitive Battery Which Included the MATRICS Consensus Cognitive Battery (MCCB) and Clinical Assessment Measures.

| **Neurocognitive Domain** | **MCCB Individual Tests** | **Citation** |
| --- | --- | --- |
| 1) Speed of Information Processing | MCCB<br>○ Brief Assessment of Cognition in Schizophrenia (BACS) Symbol Coding<br>• Category Fluency: Animal Naming<br>• Trail Making Test: Part A | Nuechterlein, 2008 |
| 2) Attention/Vigilance | MCCB<br>• Continuous Performance Test—Identical Pairs (CPT-IP) |  |
| 3) Working Memory | MCCB<br>• Wechsler Memory Scale—3rd Ed. (WMS-III): Spatial Span<br>• Letter-Number Span (LNS) |  |
| 4 Verbal Learning | MCCB<br>• Hopkin’s Verbal Learning Test—Revised (HVL-T-R) |  |
| 5) Visual Learning | MCCB<br>• Brief Visuospatial Memory Test—Revised (BVM-T-R) |  |
| 6) Reasoning and Problem Solving | MCCB<br>• Neuropsychological Assessment Battery (NAB): Mazes |  |
| 7) Social Cognition | MCCB<br>• Mayer-Salovey-Caruso Emotional Intelligence Test (MSCEIT): Managing Emotions only |  |
| **Neurocognitive Domain** | **Individual Tests** | **Citations** |
| 1) Visual Perceptual Delayed Recall | Rey–Osterrieth Complex Figure Test (ROCFT) Immediate and Delayed | Rey & Osterrieth, 1993 |
| 2) Psychomotor Functioning | Grooved Pegboard Test (dominant and non-dominant hands) | Lafayette Instrument Company, 1989 |
| 3) Executive Functioning | Trail Making Test A and B<br>Stroop Color Word Test (Stroop) | Tombaugh, 2004; Golden & Freshwater, 1978 |
| 4) Abstract Thinking | Positive and Negative Syndrome Scale (PANSS) | Kay, Fiszbein, Opler, 1987 |
| **Composite Scores** | **Individual Tests** | **Citations** |
| A. Overall Neurocognitive Composite Score | BACS: Symbol Coding, Category Fluency: Animal Naming, Trail Making Test: Part A, CPT-IP, WMS-III: Spatial Span, LNS, HVLT-R, BVM-T-R, NAB: Mazes, and MSCEIT: Managing Emotions | Nuechterlein, 2008; Rey & Osterrieth, 1993; Lafayette Instrument Company, 1989; Tombaugh, 2004; Golden & Freshwater, 1978 |
| B. Global Deficit Score (GDS) | *(A) Overall Neurocognitive Composite Score, (1) Speed of Information Processing, (2) Attention/vigilance: Continuous Performance Test—Identical Pairs (CPT-IP), (3) Working memory: Wechsler Memory Scale®—3rd Ed. (WMS®-III): Spatial Span, Letter-Number Span, (4) Verbal learning: Hopkin’s Verbal Learning Test—Revised (HVLT-R™), (5) Visual learning: Brief Visuospatial Memory Test—Revised (BVM-T-R™), (6) Reasoning and problem solving: Neuropsychological Assessment Battery® (NAB®): Mazes, (7) Social cognition: Mayer-Salovey-Caruso Emotional Intelligence Test (MSCEIT™): Managing Emotions, (8) Visual Perceptual Delayed Recall: The Rey–Osterrieth Complex Figure Test (ROCF) Immediate and Delayed, (9) Psychomotor Functioning: The Groove Pegboard (dominant and non-dominant hands), (10) Executive Functioning: Trail Making Test A and B, Stroop Color Word Test, (11) Abstract Thinking: the Positive and Negative Syndrome Scale (PANSS).* | Nuechterlein, 2008; Rey & Osterrieth, 1993; Lafayette Instrument Company, 1989; Tombaugh, 2004; Golden & Freshwater, 1978 |
| **Clinical Factors** | **Measures** | **Citation** |
| i. Depression | Beck Depression Inventory (BDI) | Beck, Steer, & Garbin, 1988 |
| ii. Sleep | Sleep Quality Assessment (PSQI) | Buyse, et al., 1989 |
| iii. Premorbid Functioning | Wechsler Test of Adult Reading (WTAR) | Wechsler, 2001 |

### 2.3. Measures

**Speed of processing, attention/vigilance, working memory, verbal learning, visual learning, reasoning and problem solving, and social cognition.** The MATRICS Consensus Cognitive Battery (MCCB; Nuechterlein et al., 2008), a paper and pencil well-validated assessment, were administered to assess seven domains of cognitive functioning.

#### 2.3.1. Visual perceptual delayed recall

The Rey–Osterrieth Complex Figure Test (ROCF; Rey & Osterrieth, 1993), a paper and pencil, well-validated measure, was utilized to assess immediate and delayed visuospatial constructional ability and visual non-verbal memory.

#### 2.3.2. Psychomotor Functioning

The Groove Pegboard (dominant and non-dominant hands; Instrument, 2002), a manipulative dexterity test, with well-established psychometric properties, was utilized to measure complex visual-motor dexterity, coordination, and performance speed.

#### 2.3.3. Executive Functioning

Trail Making Test A and B (Tombaugh, 2004), paper and pencil tasks, and the Stroop Color Word Test (Golden & Freshwater, 1978), a visual reading task, were converted to age and education corrected scores which were averaged to create a comprehensive executive functioning domain (i.e., visual searching, scanning, processing, shifting, and mental flexibility).

#### 2.3.4. Abstract Thinking

The Positive and Negative Syndrome Scale (PANSS; Kay, Fiszbein, & Opler, 1987) is usually used to measure positive and negative symptoms, and general psychopathology. The measure has well-validated reliability, criterion-related and predictive validity, within the literature analyzing individuals diagnosed with schizophrenia-spectrum disorders (Kay, Fiszbein, & Opler, 1987). We used the single PANSS item Difficulties in Abstract Thinking, which contains a section on Similarities and Proverbs to measure abstract thinking. To our knowledge this is the first time this measure has been used to assess abstract thinking in PHIV-infected-youth compared to controls.

#### 2.3.5. Developmental Level of Intellectual Functioning

To our knowledge this is the first time the Wechsler Test of Adult Reading (WTAR; Wechsler, 2001), a 10-minute reading test comprised of 50 words with irregular pronunciations, was utilized to measure estimated developmental level of intellectual functioning and quantify the impact of HIV illness on intellectual functioning prior to long-term early intervention ART intervention.

#### 2.3.6. Sleep Quality

The Sleep Quality Assessment (PSQI; Buysse et al., 1989), a self-report, well-validated measure (Backhaus et al., 2002), was utilized to measure quality and patterns of sleep. This is the first time this measure has been utilized with PHIV-infected-youth.

#### 2.3.7. Depression

The Beck Depression Inventory (BDI; Beck, Steer, & Garbin, 1988) is a common standardized 21-item, self-report measure, with high internal consistency, which has been developed to assess depressive symptomatology (Beck, Steer, & Garbin, 1988). Clinical cutoffs were determined as follows: cores of 1-10 no depression, 11-16 mild mood disturbance, 17-20 borderline clinical depression, 21-30 moderate depression, 31-40 severe depression, and over 40 extreme depression (Beck, Steer, & Garbin, 1988).

#### 2.3.8. Global Deficit Score (GDS)

A global deficit score (GDS; Carey et al., 2004). A common approach utilized to identify neurocognitive performance of adults living with HIV (Carey et al., 2004), which was calculated by converting demographically corrected t-scores from 11 neuropsychological assessment domains in addition to the Overall Neurocognitive Composite Score (see Table 2 for measures utilized to create our GDS score). A five-point scale was utilized to transform t-scores into gold standard GDS (Carey et al., 2004). To our knowledge this is the first time a GDS score has been applied to detect impairment of PHIV-infected-youth.

### 2.4. Statistical Analyses

Descriptive statistics were calculated for the demographic and neuropsychological test variables for both PHIV-infected-youth and controls. Paired t-tests were performed to determine if differences existed between PHIV-infected-youth and matched-controls. Significance level was set at 0.05 for all analyses, and the Bonferroni correction for multiple comparisons is also provided with an adjusted level of significance of 0.004. Effect size (*d*) corresponds to whether differences could be considered large between groups and was defined as d ≥ 0.80 (Sullivan & Feinn, 2012). IBM SPSS Statistics for Windows, (Version 24.0. Armonk, NY: IBM Corp. Released 2016) was utilized to perform these analyses.

## 3. Results

### Differences in neurocognitive performance between PHIV-infected youth and healthy controls

PHIV-youth performed more poorly on cognitive tasks compared to healthy controls (Table 3). Significant difference between PHIV-infected-youth and paired-healthy controls on Overall Neurocognitive Composite Score (p=0.003) and several key neurocognitive domains including Working Memory (p=0.017), Verbal Learning (p=0.000), and Attention/Vigilance (p=0.52). After correcting for multiple comparisons both Verbal Learning and Overall Neurocognitive Composite Score remained significant (p < 0.004). There were no statistically significant differences between Speed of Processing (p=.011), Attention/Vigilance (p=0.052), Reasoning and Problem Solving (p=0.211), Visual Perceptual Delayed Recall (p=0.232), and Psychomotor Functioning (p=0.114).

**Table 3.** Comparison of Neuropsychological Performance of the PHIV+ Patients (n=11 and Paired Healthy Controls (n=11).

| Variable | PHIV+<br>Mean<br>(SD) | Healthy<br>Controls<br>Mean<br>(SD) | *t* | *p* | *d* | 95%<br>Confidence<br>Interval |
| --- | --- | --- | --- | --- | --- | --- |
| Speed of Information Processing | 37.36<br>(12.56) | 51.00<br>(6.29) | 3.14 | .011* | 1.37*** | (3.95)<br>(23.33) |
| Attention/vigilance | 37.64<br>(13.96) | 48.82<br>(9.40) | 2.20 | .052 | 0.94*** | (-.14)<br>(22.50) |
| Working Memory | 37.73<br>(16.37) | 49.45<br>(7.26) | 2.85 | .017* | 0.93*** | (2.55)<br>(20.91) |
| Verbal learning | 37.55<br>(10.07) | 52.00<br>(11.30) | 5.88 | .000** | 1.35*** | (8.98)<br>(19.93) |
| Visual Learning | 38.55<br>(14.10) | 48.45<br>(5.94) | 2.65 | .024* | 0.92*** | (1.57)<br>(18.25) |
| Reasoning and Problem Solving | 41.55<br>(10.03) | 46.00<br>(9.93) | 1.34 | .211 | 0.45 | (-2.98)<br>(11.89) |
| Social Cognition | 34.00<br>(17.19) | 51.36<br>(6.77) | 3.08 | .012* | 1.33*** | (4.81)<br>(29.92) |
| Visual Perceptual Delayed Recall | 46.27<br>(13.18) | 52.27<br>(14.93) | 1.27 | .232 | 0.43 | (-4.51)<br>(16.51) |
| Psychomotor Functioning | 47.18<br>(11.61) | 52.73<br>(8.56) | 1.73 | .114 | 0.54 | (-1.58)<br>(12.67) |
| Executive Functioning | 38.36<br>(11.40) | 50.73<br>(5.82) | 3.24 | .009* | 1.37*** | (3.85)<br>(20.87) |
| Abstract Thinking | 35.00<br>(13.14) | 50.6<br>(9.63) | 3.22 | .009* | 1.36*** | (4.81)<br>(26.46) |
| Overall Neurocognitive Composite Score | 32.64<br>(15.97) | 48.91<br>(7.01) | 3.81 | .003** | 1.32*** | (6.76)<br>(25.79) |
| Global Deficit Functioning Score <sup>a</sup> | 1.37<br>(1.31) | .14<br>(.13) | -3.32 | .008* | 1.32*** | (-2.06)<br>(-.41) |

*Note.* <sup>a</sup> All scores are demographically corrected t-scores except the Global Deficit Functioning Score. The Global Deficit Score was created from gold standard conversion table (Carey et al., 2004, Arenas-Pinto et al., 2014). \* denotes significant domains $p < .05$ . \*\*denotes significant $p$
values after correction for multiple comparisons $p < .004$ . \*\*\*denotes large effect sizes in accordance with Cohen's $d$ (i.e., $d=0.2$ were considered small, $d=0.5$ medium, and $d=0.8$ large; Sullivan & Feinn, 2012.)

### Global Deficit Score (GDS)

Using Carey et al.’s (2004) GFS methodology, we found **s**ignificant differences between groups on their Global Functioning Score p=.008 indicating that PHIV youth performed more poorly compared to healthy controls (Table 3).

### Comparisons between groups on assessments of sleep and depression

Significant differences between groups were noted for self-reported depressive symptoms (*p* < 0.047). The PHIV-infected-youth (M=3.09) reported significantly higher levels of depression (9% reported moderate depression, 18% reported borderline clinical depression, 9% mild mood disturbance, and 63% reported no depression) than the healthy control group (100% of reported ups and downs are considered within normal limits). No significant differences were present under self-reported sleep quality.

### Developmental level of intellectual functioning between group comparison

There were significant differences between groups on estimated developmental level of intellectual functioning *p* < 0.001. The PHIV-infected-youth performed significantly worse than the healthy control group when predicting neurocognitive and memory functioning prior to illness progression as measured by the WTAR. These results are consistent with early onset of central nervous system pathology.

## 4. Discussion

Consistent with prior studies, our results demonstrate the existence of neurocognitive deficits between PHIV-infected-youth with early, prolonged, and current antiretroviral treatment and the paired healthy age and gender matched control group. Specifically, significant neurocognitive performance deficits emerged between groups on Overall MCCB Neurocognitive Composite Scores (p=0.003), Working Memory, Verbal Learning, Visual Learning, Social Cognition, Executive Functioning, Abstract Thinking, and a Global Deficit Score. After correction for multiple comparisons, both Verbal Learning and the Overall Neurocognitive Composite Score differences remained statistically significant (p<0.004). Our study corroborates previous findings that individuals diagnosed with PHIV have neurocognitive impairments (Cohen et al., 2014; Hermetet-Lindsay et al., 2017; Laughton et al., 2013; Malee, Smith, Mellins et al., 2016; Nagarajan et al., 2012; Nichols et al., 2015; Sarma et al., 2014; Sirois et al., 2016; Willen et al., 2017; Yadav et al., 2017). These deficiencies persist into adulthood despite antiretroviral therapy and most likely reflect the early infection of the central nervous system. Furthermore, additional support for neurocognitive deficits comes from the very large effect sizes for Speed of Information Processing, Verbal Learning, Social Cognition, Executive Functioning, Abstract Thinking, Overall Neurocognitive Composite Score, and Global Deficit Scores.

Our research indicated this is the first time a GDS score has been applied to detect neurocognitive impairments of PHIV-infected-youth. Although the GDS did not meet the multiple comparisons cutoff (Table 3) there was a large effect size which displayed significant differences between groups (*d*=1.32). Laughton et al. (2013), noted that global cognitive scores might overlook subtle individual domain impairments. A strength of the present study is that we utilized a comprehensive battery of assessments to better assess each neurocognitive domain and the findings are so strikingly large that it’s unlikely that a larger sample size would alter the current findings. Furthermore, to our knowledge this is the first study to examine sleep, abstract thinking, and developmental level of intellectual functioning to measure estimated early intellectual functioning and quantify the impact of HIV illness on intellectual functioning.

Our results suggest there are no significant differences between groups on Attention/Vigilance, Reasoning and Problem Solving, Visual Perceptual Delayed Recall, or Psychomotor Functioning. Nonetheless, our sample size is small and previous studies have reported differences in some of these domains. Cohen et al. (2014), found children (8 to 18-years-old) with PHIV performed worse on all neurocognitive domains assessed (i.e., Verbal IQ, Performance IQ, Processing Speed, Working Memory, Motor Functioning, Memory, and Executive Functioning) than controls. Furthermore, we also previously found a deficit in Attention / Processing Speed in 13 to 25-year-old adolescent PHIV-infected-youth^2^. Some researchers (Laughton et al., 2013; Nichols et al., 2015; Willen et al., 2017) also found a significant difference between PHIV-infected-youth and matched healthy controls on measures of Executive Functioning; however, others (Laughton et al., 2013; Willen et al., 2017;) including our study, did not find significant differences on measures of Psychomotor Speed. Recently, Van den Hof et al. (2019), reported a neurocognitive longitudinal study of the NOVICE cohort PHIV as they progressed into adolescence over 4.6 years and demonstrated worsening executive function over time, while Robbins et al. (2019), evaluated children as they progressed into adulthood longitudinally and noted better performance on both Executive Functioning and Working Memory over time. Their results suggest the need for further studies to assess neurocognitive issues as PHIV-youth mature.

The verbal learning deficit was striking for our PHIV young adults who received prolonged and early treatment for HIV. With a p value of 0.000, the difference was still significant after correction for multiple comparisons. The Verbal Learning domain was one of only two domains for which the standard deviation was smaller for PHIV-infected-youth compared to controls, which supports how common the deficit in verbal learning is in our PHIV patients. For most domains, the standard deviation was higher for the patients than controls suggesting much more variation in the PHIV-infected-youth group. A confounding variable for our study could be the effect on language development of living within a bilingual environment or children born in other countries. We did not track this information since all of the participants were fluent in English. Others have also reported verbal impairments including Cohen et al. (2014), who studied PHIV in early adolescence (median age 13.8 years, p=0.043). Sirois et al. (2016), looked at associations of memory and executive functioning with academic and adaptive functioning. Verbal learning was statistically associated with measures of academic achievement at p<0.001 in PHIV. Adults who acquired HIV have also been noted to have verbal memory impairments (Wright et al., 2011). PHIV-youth performed significantly worse than healthy controls on developmental level of intellectual functioning, memory which were likely present prior to long-term early intervention ART intervention WTAR (*t*(10) = 4.357, *p* < .001). Impairments in intellectual functioning have also been noted in the literature for individuals with PHIV prior to ART intervention (Laughton et al., 2013). Therefore, we believe that these impairments are part of the condition of HIV and thus chose not to treat developmental level of intellectual functioning (WTAR) as a covariate in our analysis.

Similar to our study, Ireland (2011), compared a small sample of individuals diagnosed with HIV-Associated Neurocognitive Disorders (HAND) to controls and found early impairments in Social Cognition regardless of functioning on other cognitive domains. Louthrenoo, Oberdorfer, and Sirisanthana (2014), also found that adolescents with PHIV had higher social withdrawal compared to controls. In furtherance of these results, we examined difference in ethnicity (p-value = 0.086) and education (p-value = 0.476) with the results of the Fisher’s exact test, and found no significant difference between groups; thus, we concluded language development was equivalent across both groups. Deficiency in Verbal Learning and Social Cognition might have a significant impact on physician/patient interactions and enhanced awareness of this deficiency would be important to improve medical communication between patients and physicians. Morgan et al. (2019), also noted poorer healthcare interactions between those diagnosed with HAND compared to controls.

Recent studies have shown that other risk factors such as stressful life events, socioeconomic status (Malee et al., 2016; Mellins & Malee, 2013; Nichols et al., 2015; Smith & Wilkins, 2015), urban violence (Smith & Wilkins, 2015; Willen et al., 2017), disadvantaged neighborhoods (Abrams et al., 2018; Malee et al., 2016; Mellins & Malee, 2013), objective perception of their neighborhoods (Kang et al., 2019), familial instability (Smith & Wilkins, 2015), family history of mental illness (Mellins & Malee, 2013; Nichols et al., 2015), individual health status (Mellins & Malee, et al., 2013; Smith et al., 2019), low educational enrichment (Nichols et al., 2015), and the stigmatizing nature of HIV (Abrams et al., 2018; Smith et al., 2019; Willen et al., 2017) may also relate to neurocognitive deficits and mental health outcomes. Smith et al. (2019), reported that when PHIV youth were compared to HIV-exposed but uninfected youth, 36% of the youth had a previous or current mental health diagnosis with no significant HIV status group differences. Due to these potential risk factors individuals diagnosed with PHIV may have limited access to educational enrichment opportunities and these differences may also act as a potential confound when reporting neuropsychological outcomes.

Although there was no evidence of sleep difficulties, our PHIV-infected-youth were significantly more depressed than the controls, even though none of the patients were actively receiving psychiatric care for depression. These results are consistent with other studies of this population (Hermetet-Lindsay et al., 2017; Malee et al., 2011; Mellins & Malee, 2013; Smith et al., 2019; Vreeman et al., 2015) which suggest that undiagnosed depression could be an important issue of care for these patients. Le Prevost et al. (2018), showed that there were no differences in anxiety and depression between PHIV and control participants in an adolescent population. Depression and anxiety appear to be more prevalent than is clinically recognized. However, research suggests that co-morbid mental health diagnoses are common, and may not be related to HIV-infection status (Smith et al., 2019). Additionally, parent support decreases depression, as well as increases cognitive development (Laughton et al., 2013). Due to increased risk of mental health diagnoses, Smith et al. (2019) have recommended multi-faceted approaches that incorporate both individual and family participation in treatment programs.

## 5. Study Limitations

There are a number of limitations that should be considered when interpreting the generalizability of our results including our relatively small sample size (Sarma et al., 2021). Our study did not assess participants’ bilingual status, level of acculturation, or sociodemographic factors, which could confound our findings. Furthermore, we were also unable to match for race; however, our results did show that race was not deemed significantly pertinent to the results of this study. Although our results indicate PHIV-participants differ from our control group on various neurocognitive factors, these results may be less generalizable due to the potential confounding variables listed above.

## 6. Conclusions

Findings show that despite long-term cART treatment, neurocognitive deficiencies remain for PHIV-infected-youth into adulthood. Although the majority of our PHIV (7/11) were treated before the first birthday, even earlier treatment could enhance the neuropsychiatric and neurocognitive outcomes by aborting development of HIV encephalopathy or subclinical involvement of the central nervous system. The deficiency in the Verbal Learning domain was highly significant and withstood correction for multiple comparisons. Awareness of this defect could improve communication between patients and their medical care team. Furthermore, mental health diagnoses such as depression should be considered and addressed routinely.

## Data Availability

All data produced in the present study are available upon reasonable request to the authors.

## 7. Acknowledgements

This research was supported by a research grant from the National Institute of Neurological Disorders and Stroke (NINDS) [R21NS090956].

## Ethics & Consent to participate & publish

Informed, signed consent was obtained from each subject using an Institutional Review Board (IRB) consent form approved by the Los Angeles Biomedical Research Institute at Los Angeles County Harbor-UCLA Medical Center (now Lundquist Institute). Both the Los Angeles Biomedical Research Institute and UCLA gave IRB approval which also included MRI/MRS imaging, neurocognitive assessment, and a detailed review of the patient subjects’ medical records (see Sarma et al., 2021).

## Data Availability Statement

The authors do not have permission to share raw subject data.

## Code Availability

The authors are able to share methods of coding and analyzing data

## Authors’ contributions

TW, MK, MKS, JS, JV, and MAT, contributed to the design of design and the conduct of the study. TW, MK, MKS, JH, JV, and MAT, contributed to data collection. TW, MK, MKS, IW, DEM, JH, KN, JD, AK, EO, JAC, MAT, and JV, contributed to patient-related coordination and management of recruitment and enrollment. TW, MK, JS, and JV contributed to data analysis and interpretation. All authors contributed to writing the manuscript.

## References

1. Abrams, E. J., Mellins, C. A., Bucek, A., Dolezal, C., Raymond, J., Wiznia, A., … & Ng, Y. K. W. (2018). Behavioral health and adult milestones in young adults with perinatal HIV infection or exposure. Pediatrics, 142(3). doi: 10.1542/peds.2018-0938.

2. Ashby, J., Foster, C., Garvey, L., Wan, T., Allsop, J., Paramesparan, Y., … & Winston, A. (2015). Cerebral function in perinatally HIV-infected young adults and their HIV-uninfected sibling controls. HIV clinical trials, 16(2), 81–87. doi: 10.1179/1528433614Z.0000000003.

3. Backhaus, J., Junghanns, K., Broocks, A., Riemann, D., & Hohagen, F. (2002). Test– retest reliability and validity of the Pittsburgh Sleep Quality Index in primary insomnia. Journal of psychosomatic research, 53(3), 737–740. doi: 10.1016/S0022-3999(02)00330-6.

4. Basso, M. R., & Bornstein, R. A. (2000). Estimated premorbid intelligence mediates neurobehavioral change in individuals infected with HIV across 12 months. Journal of clinical and experimental neuropsychology, 22(2), 208–218. doi: 10.1076/1380-3395(200004)22:2;1-1;FT208.

5. Beck, A. T., Steer, R.A., & Garbin, M.G. (1988) Psychometric properties of the Beck Depression Inventory: Twenty-five years of evaluation. Clinical psychology review, 8(1), 77–100. doi: 10.1016/0272-7358(88)90050-5.

6. Buysse, D. J., Reynolds III, C. F., Monk, T. H., Berman, S. R., & Kupfer, D. J. (1989). The Pittsburgh Sleep Quality Index: a new instrument for psychiatric practice and research. Psychiatry research, 28(2), 193–213. doi: 10.1016/0165-1781(89)90047-4.

7. Carey, C. L., Woods, S. P., Gonzalez, R., Conover, E., Marcotte, T. D., Grant, I., & Heaton, R. K. (2004). Predictive validity of global deficit scores in detecting neuropsychological impairment in HIV infection. Journal of clinical and experimental neuropsychology, 26(3), 307–319. doi: 10.1080/13803390490510031.

8. Cohen, S., Ter Stege, J. A., Geurtsen, G. J., Scherpbier, H. J., Kuijpers, T. W., Reiss, P., … & Pajkrt, D. (2014). Poorer cognitive performance in perinatally HIV-infected children versus healthy socioeconomically matched controls. Clinical infectious diseases, 60(7), 1111–1119. doi: 10.1093/cid/ciu1144

9. Chernoff, M., Angelidou, K. (Nadia), Williams, P. L., Brouwers, P., Warshaw, M., & Nachman, S. (2018). Assessing psychiatric symptoms in youth affected by HIV: comparing a brief self-administered rating scale with a structured diagnostic interview. Journal of clinical psychology in Medical settings, 25(4), 420–428. doi: 10.1007/s10880018-9550-2.

10. Golden, C. J., & Freshwater, S. M. (1978). Stroop color and word test: a manual for clinical and experimental uses. Chicago: Stoelting.

11. Hermetet-Lindsay, K. D., Correia, K. F., Williams, P. L., Smith, R., Malee, K. M., Mellins, C. A., & Rutstein, R. M. (2017). Contributions of disease severity, psychosocial factors, and cognition to behavioral functioning in US youth perinatally exposed to HIV. AIDS and Behavior, 21(9), 2703–2715. doi: 10.1007/s10461-016-1508-5.

12. Instrument, L. (2002). Grooved Pegboard test user instructions. Lafayette: Lafayette Instrument.

13. Ireland, E. (2011). Exploring social cognition and executive function in HIV-Associated Neurocognitive Disorders (HAND). (Doctoral dissertation, University of East London), UK.

14. Kang, E., Leu, C. S., Snyder, J., Robbins, R. N., Bucek, A., & Mellins, C. A. (2019). Caregiver perceptions of environment moderate relationship between neighborhood characteristics and language skills among youth living with perinatal HIV and uninfected youth exposed to HIV in New York City. AIDS care, 31(1), 61–68. doi: 10.1080/09540121.2018.1492698.

15. Kay, S. R., Fiszbein, A., & Opler, L. A. (1987). The positive and negative syndrome scale (PANSS) for schizophrenia. Schizophrenia bulletin, 13(2), 261–276. doi: 10.1093/schbul/13.2.261.

16. Laughton, B., Cornell, M., Boivin, M., & Van Rie, A. (2013). Neurodevelopment in perinatally HIV infected children: a concern for adolescence. Journal of the International AIDS Society, 16(1), 18603. doi: 10.7448/IAS.16.1.18603.

17. Le Prevost, M., Arenas-Pinto, A., Melvin, D., Parrott, F., Foster, C., Ford, D., … & Gibb, D. M. (2018). Anxiety and depression symptoms in young people with perinatally acquired HIV and HIV affected young people in England. AIDS Care - Psychological and Socio-Medical Aspects of AIDS/HIV, 30(8), 1040–1049. doi: 10.1080/09540121.2018.1441972.

18. Louthrenoo, O., Oberdorfer, P., & Sirisanthana, V. (2014). Psychosocial functioning in adolescents with perinatal HIV infection receiving highly active antiretroviral therapy. Journal of the International Association of Providers of AIDS Care, 13(2), 178–183. doi: 10.1177/2325957413488171.

19. Malee, K. M., Smith, R. A., & Mellins, C. A. (2016). Brain and cognitive development among US youth with perinatally acquired human immunodeficiency virus infection. Journal of the Pediatric Infectious Diseases Society, 5(Suppl_1), S1–S5. doi: 10.1093/jpids/piw041.

20. Malee, K. M., Tassiopoulos, K., Huo, Y., Siberry, G., Williams, P. L., Hazra, R., … & Kapetanovic, S. (2011). Mental health functioning among children and adolescents with perinatal HIV infection and perinatal HIV exposure. AIDS Care - Psychological and Socio-Medical Aspects of AIDS/HIV, 23(12), 1533–1544. doi: 10.1080/09540121.2011.575120.

21. Mellins, C. A., & Malee, K. M. (2013). Understanding the mental health of youth living with perinatal HIV infection: lessons learned and current challenges. Journal of the International AIDS Society, 16(1). doi: 10.7448/IAS.16.1.18593.

22. Morgan, E. E., Woods, S. P., Iudicello, J. E., Grant, I., Villalobos, J., & HIV Neurobehavioral Research Program (HNRP) Group. (2019). Poor self-efficacy for healthcare provider interactions among individuals with HIV-associated neurocognitive disorders. Journal of clinical psychology in medical settings, 26(1), 13–24. doi: 10.1007/s10880-018-9560-0.

23. Nagarajan, R., Sarma, M. K., Thomas, M. A., Chang, L., Natha, U., Wright, M., … & Keller, M. A. (2012). Neuropsychological function and cerebral metabolites in HIV-infected youth. Journal of Neuroimmune Pharmacology, 7(4), 981–990. doi: 10.1007/s11481-012-9407-7.

24. Nichols, S. L., Brummel, S. S., Smith, R. A., Garvie, P. A., Hunter, S. J., Malee, K. M., … Pediatric HIVAIDS Cohort Study (2015). Executive Functioning in Children and Adolescents With Perinatal HIV Infection. The Pediatric infectious disease journal, 34(9), 969–75. doi: 10.1097/INF.0000000000000809.

25. Nuechterlein, K. H., Green, M. F., Kern, R. S., Baade, L. E., Barch, D. M., Cohen, J. D., … & Goldberg, T. 2008). The MATRICS Consensus Cognitive Battery, part 1: test selection, reliability, and validity. American Journal of Psychiatry, 165(2), 203–213. doi: 10.1176/appi.ajp.2007.07010042.

26. Rey, A., & Osterrieth, P. A. (1993). Translations of excerpts from Andre Rey’ s Psychological examination of traumatic encephalopathy and PA Osterrieth’s The Complex Figure Copy Test. Clinical Neuropsychologist.

27. Robbins, R. N., Zimmerman, R., Korich, R., Raymond, J., Dolezal, C., Choi, C. J., … & Abrams, E. J. (2019). Longitudinal trajectories of neurocognitive test performance among individuals with perinatal HIV-infection and-exposure: adolescence through young adulthood. AIDS Care - Psychological and Socio-Medical Aspects of AIDS/HIV, 1–9. doi: 10.1080/09540121.2019.1626343.

28. Sarma, M. K., Nagarajan, R., Keller, M. A., Kumar, R., Nielsen-Saines, K., Michalik, D. E., … & Thomas, M. A. (2014). Regional brain gray and white matter changes in perinatally HIV-infected adolescents. NeuroImage: Clinical, 4, 29–34. doi: 10.1016/j.nicl.2013.10.012.

29. Sarma, K M., Pal, A., Keller, M., Welikson, T., Ventura, J., Michalik, E D., Nielsen-Saines, K., Deville., J, Kovacs, A., Operskalski, E., Church, A J., Macey, M. M., Biswal, B., Thomas, M A. (2021). White matter of perinatally HIV infected older youths shows low frequency fluctuations that may reflect glial cycling. Scientific Reports. doi: 10.1038/s41598-02182587-5

30. Sirois, P. A., Chernoff, M. C., Malee, K. M., Garvie, P. A., Harris, L. L., Williams, P. L., … & Nichols, S. L. (2016). Associations of memory and executive functioning with academic and adaptive functioning among youth with perinatal HIV exposure and/or infection. Journal of the Pediatric Infectious Diseases Society, 5(Suppl_1), S24–S32. doi: 10.1093/jpids/piw046.

31. Smith, R., Huo, Y., Tassiopoulos, K., Rutstein, R., Kapetanovic, S., Mellins, C., … & Pediatric HIV/AIDS Cohort Study (PHACS). (2019). Mental health diagnoses, symptoms, and service utilization in US youth with perinatal HIV Infection or HIV Exposure. AIDS patient care and STDs, 33(1), 1–13. doi: 10.1089/apc.2018.0096.

32. Smith, R., & Wilkins, M. (2015). Perinatally acquired HIV infection: long-term neuropsychological consequences and challenges ahead. Child Neuropsychology, 21(2), 234–268. doi: 10.1080/09297049.2014.898744.

33. Sullivan, G. M., & Feinn, R. (2012). Using effect size—or why the P value is not enough. Journal of graduate medical education, 4(3), 279–282. doi: 10.4300/jgme-d-12-00156.1.

34. Tombaugh, T. N. (2004). Trail Making Test A and B: normative data stratified by age and education. Archives of clinical neuropsychology, 19(2), 203–214. doi: 10.1016/S0887-6177(03)00039-8.

35. Van den Hof, M., Ter Haar, A. M., Scherpbier, H. J., van der Lee, J. H., Reiss, P., Wit, F. W., … & Pajkrt, D. (2019). Neurocognitive development in perinatally HIV-infected adolescents on long-term treatment compared to healthy matched controls: a longitudinal study. Clinical Infectious Diseases. doi: 10.1093/cid/ciz386.

36. Vreeman, R. C., Scanlon, M. L., McHenry, M. S., & Nyandiko, W. M. (2015). The physical and psychological effects of HIV infection and its treatment on perinatally HIV infected children. Journal of the International AIDS Society, 18(Suppl_6), 20258. doi: 10.7448/IAS.18.7.20258.

37. Wechsler, D. (2001). Wechsler Test of Adult Reading: WTAR. San Antonio, TX: NCS Pearson.

38. Willen, E. J., Cuadra, A., Arheart, K. L., Post, M. D., & Govind, V. (2017). Young adults perinatally infected with HIV perform more poorly on measures of executive functioning and motor speed than ethnically matched healthy controls. AIDS Care - Psychological and Socio-Medical Aspects of AIDS/HIV, 29(3), 387–393. doi:10.1080/09540121.2016.1234677.

39. Wright, M. J., Woo, E., Foley, J., Ettenhofer, M. L., Cottingham, M. E., Gooding, A. L., … & Hinkin, C. H. (2011). Antiretroviral adherence and the nature of HIV-associated verbal memory impairment. The Journal of neuropsychiatry and clinical neurosciences, 23(3), 324–331. doi: 10.1176/jnp.213.jnp324.

40. Yadav, S. K., Gupta, R. K., Garg, R. K., Venkatesh, V., Gupta, P. K., Singh, A. K., & Marincola, F. M. (2017). Altered structural brain changes and neurocognitive performance in pediatric HIV. NeuroImage: Clinical, 14, 316. 10.1016/j.nicl.2017.01.032.

